# Clinical characteristics and outcomes of inpatients with neurological disease and COVID-19

**DOI:** 10.1101/2020.04.28.20082735

**Authors:** Alberto Benussi, Andrea Pilotto, Enrico Premi, Ilenia Libri, Marcello Giunta, Chiara Agosti, Antonella Alberici, Enrico Baldelli, Matteo Benini, Sonia Bonacina, Laura Brambilla, Salvatore Caratozzolo, Matteo Cortinovis, Angelo Costa, Stefano Cotti Piccinelli, Elisabetta Cottini, Viviana Cristillo, Ilenia Delrio, Massimiliano Filosto, Massimo Gamba, Stefano Gazzina, Nicola Gilberti, Stefano Gipponi, Alberto Imarisio, Paolo Invernizzi, Ugo Leggio, Matilde Leonardi, Paolo Liberini, Martina Locatelli, Stefano Masciocchi, Loris Poli, Renata Rao, Barbara Risi, Luca Rozzini, Andrea Scalvini, Francesca Schiano di Cola, Raffaella Spezi, Veronica Vergani, Irene Volonghi, Nicola Zoppi, Barbara Borroni, Mauro Magoni, Alessandro Pezzini, Alessandro Padovani

## Abstract

**Objective:** To report the clinical and laboratory characteristics, as well as treatment and clinical outcomes of patients admitted for neurological diseases with and without COVID-19.

**Methods:** In this retrospective, single center cohort study, we included all adult inpatients with confirmed COVID-19, admitted to a Neuro-COVID Unit from February 21, 2020, who had been discharged or died by April 5, 2020. Demographic, clinical, treatment, and laboratory data were extracted from medical records and compared (FDR-corrected) to those of neurological patients without COVID-19 admitted in the same period.

**Results:** 173 patients were included in this study, of whom 56 were positive for COVID-19 while 117 were negative for COVID-19. Patients with COVID-19 were older, had a different distribution regarding admission diagnoses, including cerebrovascular disorders, and had a higher quick Sequential Organ Failure Assessment (qSOFA) score on admission (all *p*<0.05). In-hospital mortality rates and incident delirium were significantly higher in the COVID-19 group (all *p*<0.005). COVID-19 and non-COVID patients with stroke had similar baseline characteristics but patients with COVID-19 had higher modified Rankin scale scores at discharge, with a significantly lower number of patients with a good outcome (all *p*<0.001).

In patients with COVID-19, multivariable regressions showed increasing odds of in-hospital death associated with higher qSOFA scores (odds ratio 4.47, 95% CI 1.21-16.5; *p*=0.025), lower platelet count (0.98, 0.97-0.99; *p*=0.005) and higher lactate dehydrogenase (1.01, 1.00-1.03; *p*=0.009) on admission.

**Conclusions:** COVID-19 patients admitted with neurological disease, including stroke, have a significantly higher in-hospital mortality, incident delirium and higher disability than patients without COVID-19.

## Introduction

Since February 20, 2020, Lombardy, Italy, has experienced a major outbreak of coronavirus disease 2019 (COVID-19), caused by the severe acute respiratory syndrome coronavirus 2 (SARS-CoV-2), with more than 50,000 cases and 9,000 deaths in the region as of April 5, 2020.^1^

The clinical spectrum of SARS-CoV-2 appears to be wide, encompassing asymptomatic infections, mild upper respiratory tract illness and severe pneumonia with respiratory failure and death.^2^ Several factors have been associated with increased mortality, including older age, high Sequential Organ Failure Assessment (SOFA) score, and increased d-dimer levels.^3^

To date, only two retrospective case series from convenience samples from three hospitals in Wuhan, China, have been published^4^ or posted on pre-print servers without peer review.^5^ The most common neurological manifestations were dizziness (16.8%), headache (13.1%) and encephalopathy (2.8%). Stroke complicated COVID-19 infection in 5.9% of cases, with patients being older, with more cardiovascular comorbidities, and more severe pneumonia.^5,6^

It is yet unclear if patients with neurological disease and COVID-19 have a different neurological outcome compared to patients without COVID-19, and if this is achieved at the cost of days of hospitalization or increased mortality. Moreover, it is not known if stroke severity at admission and at discharge are similar in the two populations, and if acute phase treatments, as endovascular therapy and intravenous fibrinolysis have similar outcomes. We aimed to describe the clinical and laboratory characteristics, as well as treatment and clinical outcomes of patients with neurological disease, with and without COVID-19.

## Methods

### Standard Protocol Approvals, Registrations, and Patient Consents

This study received approval from the ethical standards committee on human experimentation (local ethics committee of the ASST Spedali Civili Hospital, Brescia: NP 4051, approved April 6, 2020). The requirement for informed consent was waived by the ethics commission for patients who were not longer alive or reachable at the time of approval, while full written informed consent was required for all other participants.

### Study design and participants

This retrospective cohort study included adult inpatients (≥18 years old), admitted primarily for neurological disease, from the General Neurology Unit and Vascular Neurology Unit, Department of Neurological and Vision Sciences, ASST Spedali Civili Hospital, Brescia, Italy from February 21 to April 5, 2020. This Hospital in the City of Brescia was designated as a “hub” for acute cerebrovascular diseases during the COVID-19 outbreak, in a metropolitan area of more than 1,200,000 people.^7^ The original units were divided in two separate sections for patients affected (Neuro-COVID unit) and non-affected (non-COVID unit) by COVID-19, and staff neurologists were equally divided between the two units.^8^

Our study included all adult inpatients who were hospitalized for neurological diseases and had a definite outcome (discharge home or to a rehabilitation facility, or death). The criteria for discharge for patients with COVID-19 were absence of fever for at least 24 hours, a respiratory rate <22/min, and substantial improvement at chest x-ray or computed tomography (CT) scan.

### Data collection

Epidemiological, demographical, clinical, laboratory, treatment, and outcome data were extracted from both printed and electronic medical records using standardized anonymized data collection forms. All data were imputed and checked by four physicians (AB, AP, MG and IL). The admission data of included patients ranged from February 21 to April 5, 2020.

### Demographical and clinical data

The following demographic and clinical data were acquired for all patients, which were present on admission: age, sex, smoking habits, comorbidities (diabetes, hypercholesterolemia, hypertension, coronary heart disease, malignancy, chronic kidney disease, immunodeficiency), the quick Sequential Organ Failure Assessment (qSOFA) score, the premorbid modified Rankin score (mRS), the National Institute of Health Stroke Scale (NIHSS) score (for cerebrovascular disease only); during hospitalization: antibiotic, antiviral or high-flow-oxygen therapy, in-hospital mortality, incident delirium, fever during hospitalization, number of diagnostic tests, acute phase therapies as intravenous fibrinolysis, endovascular therapy or bridging therapy (for ischemic stroke only); or at discharge: days of hospitalization, mRS, NIHSS score (for cerebrovascular disease only). The qSOFA score uses three criteria, assigning one point for low systolic blood pressure (≤100 mmHg), high respiratory rate (≥22 breaths per minute), or altered mentation (Glasgow coma scale<15), with a range from 0 (least impairment) to 3 (greatest impairment).

### Laboratory procedures

SARS-CoV-2 detection in respiratory specimens was performed by real-time RT PCR methods, as described elsewhere.^9^ Both nasopharyngeal and oropharyngeal swabs were performed in all patients. If two consecutive tests obtained at least 24 hours apart resulted negative, and there was high suspicion of COVID-19 (i.e. interstitial pneumonia at chest x-ray, low arterial partial pressure of oxygen), a bronchoalveolar lavage was performed.

Routine blood examinations comprised complete blood count, erythrocyte sedimentation rate, serum biochemical tests including C-reactive protein, liver and renal function, lactate dehydrogenase, creatine kinase, high-sensitivity troponin T, serum ferritin and coagulation profile. High sensitivity troponin T, ferritin and d-dimer were performed only in a subset of patients (~20%).

### Definitions

Fever was defined as axillary temperature of at least 37.5°C. The diagnosis of delirium was defined by the presence of features 1 and 2 and either 3 or 4 at the Confusion Assessment Method (CAM).^10^ Antiviral treatment was defined as lopinavir/ritonavir 200/50 mg 2 cp BID, darunavir 800 mg 1 cp QD + ritonavir 100 mg 1 cp QD, or darunavir/cobicistat 800/150 mg 1 cp QD. In stroke patients, a good outcome was defined as a mRS score ≤2. Diagnostics tests were defined as magnetic resonance imaging (head), CT (head/thorax/abdomen), X-ray (thorax/abdomen), electroencephalography, echography (heart/neck), and Holter monitor.

### Statistical analysis

Continuous and categorical variables are reported as median (interquartile range) and n (%) respectively. Differences between patients with and without COVID-19 were compared by Mann-Whitney U test, χ^2^ test or Fisher’s exact test where appropriate.

To explore the risk factors associated with in-hospital death, univariable and multivariable logistic regression models were implemented. For multivariate analysis, to avoid overfitting in the model, variables were chosen based on previous findings and clinical constraints.^3^ Previous studies have shown age, qSOFA scores and several laboratory findings to be associated with in-hospital mortality. Therefore, we chose age, qSOFA scores, platelet count, C-reactive protein and lactate dehydrogenase for our multivariable logistic regression model.

A two-sided *p*-value<0.05 was considered significant and corrected for multiple comparisons using the Benjamini-Hochberg false discovery rate (FDR).^11^ Data analyses were carried out using SPSS software (version 21.0) and GraphPad Prism (version 8.0).

### Data availability

All study data, including raw and analyzed data, and materials will be available from the corresponding author, A.P., upon reasonable request.

## Results

Two hundred fourteen adult patients were hospitalized in the Neurology and Vascular Neurology Unit of the ASST Spedali Civili di Brescia Hospital from February 21 to April 5, 2020. After excluding 41 patients which were still hospitalized as of April 5, 2020, we included 173 inpatients in the final analysis. Of these, 56 (32.4%) resulted positive for COVID-19 and were admitted in the Neuro-COVID unit (see **Fig. 1**).

**Figure 1.**
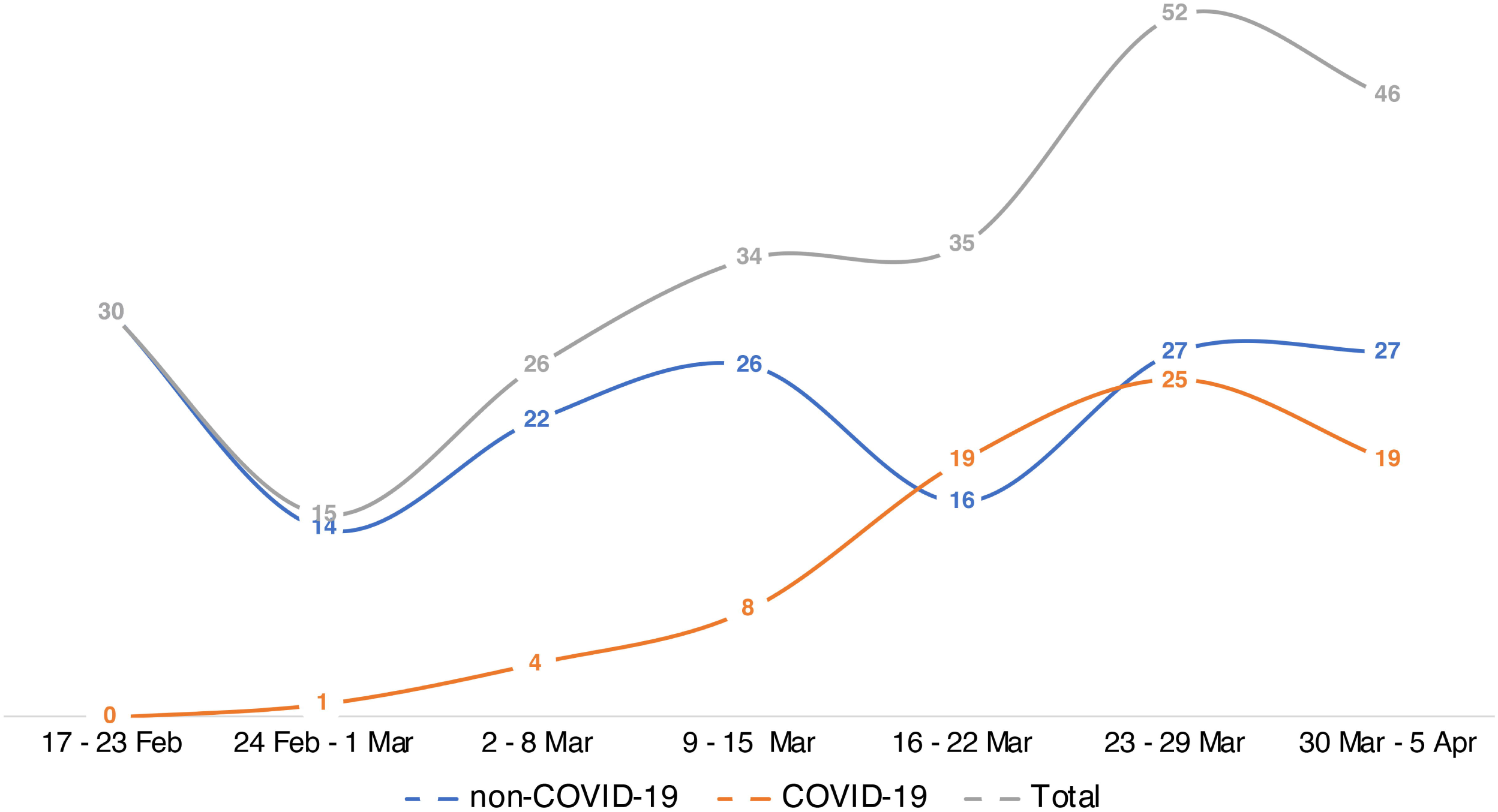
Weekly admissions of patients with neurological diseases, with and without COVID-19.

Demographic, clinical and laboratory characteristics of included patients are reported in **Table 1**. Results are reported as median (IQR) or n (%), while FDR-adjusted *p*-values for multiple comparisons are reported for each test. Compared to patients without COVID-19, patients with COVID-19 were older (77.0, IQR 67.0-83.8 vs 70.1, IQR 52.9-78.6, *p*=0.006), had a different distribution regarding admission diagnoses particularly for cerebrovascular disorders (n=43, 76.8% vs n=68, 58.1%, *p*=0.035), and had a higher qSOFA score on admission (0.5, IQR 0.4-0.6 vs 0.9, IQR 0.7-1.1, *p*=0.006). No significant differences were observed for comorbidities including diabetes, hypercholesterolemia, hypertension, coronary heart disease, chronic kidney disease, immunodeficiency or malignancy (all *p*>0.050). COVID-19 patients had higher in-hospital mortality (n=21, 37.5% vs n=5, 4.3%, *p*<0.001), higher incidence of delirium (n=15, 26.8% vs n=9, 7.7%, *p*=0.003) and fever (n=27, 48.2% vs n=14, 12.0%, *p*<0.001), while days of hospitalization were similar (n=6.0 IQR 3.3-10.0 vs 5.0, IQR 4.0-8.0, *p*=0.424). Of patients that were discharged (excluding in-hospital deaths), days of hospitalization were increased in COVID-19 patients (8.0, IQR 5.0-11.0 vs 5.0, IQR 4.0-8.0, *p*=0.005).

**Table 1.**
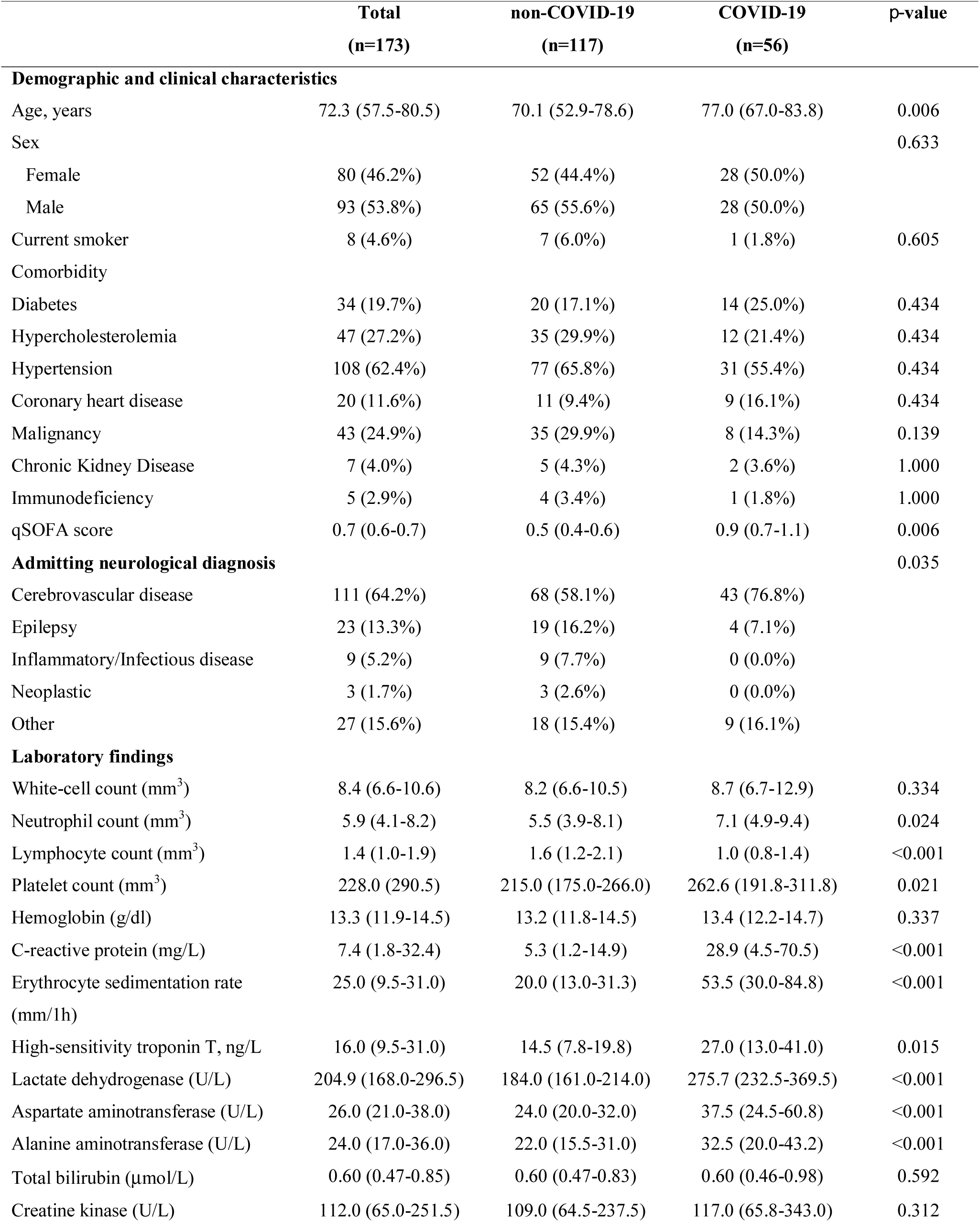
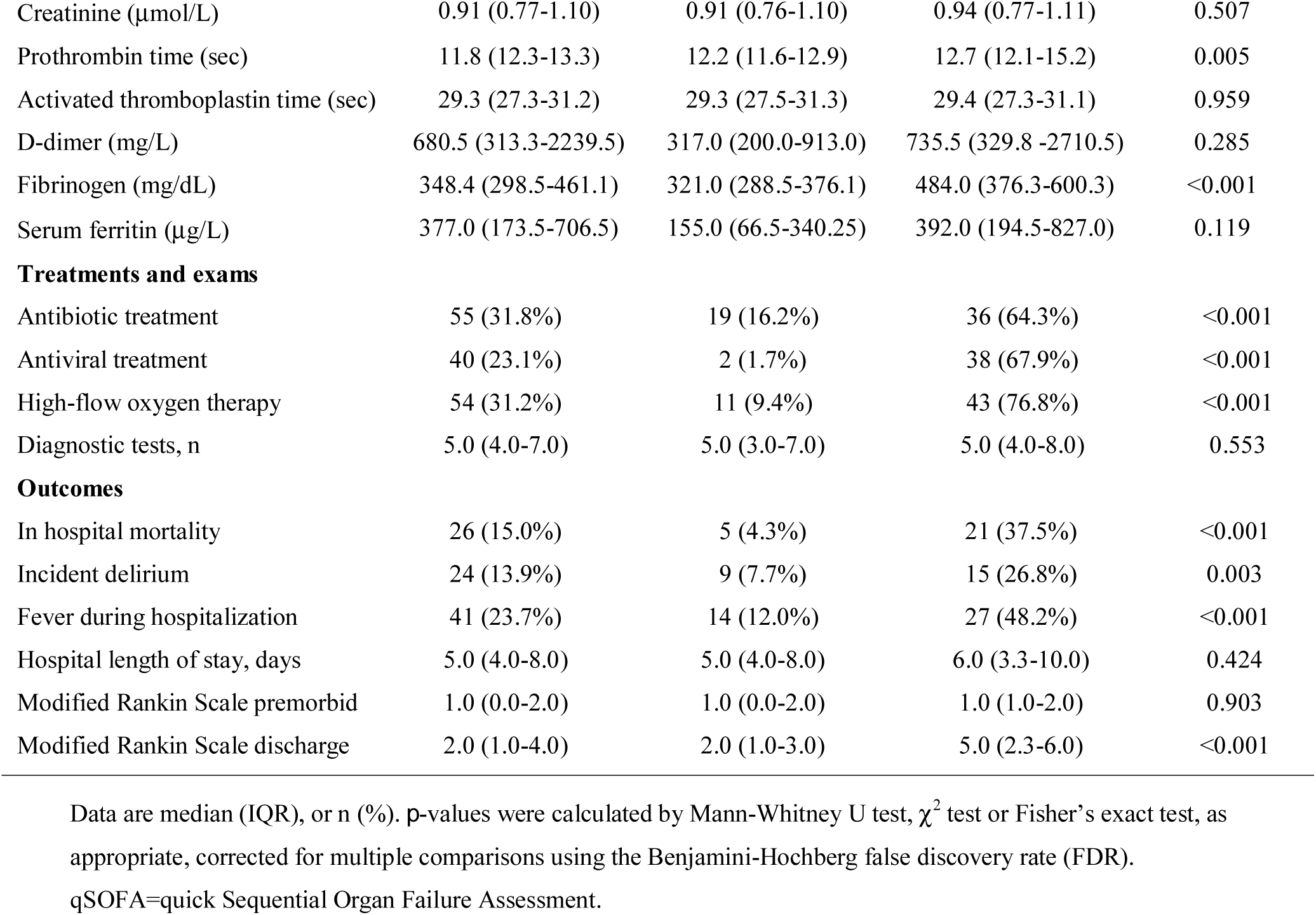
Demographic, clinical, laboratory characteristics, treatment and clinical outcomes of all included patients.

Treatments were different between the two groups, with a wider use of high flow oxygenation (n=43, 76.8% vs n=11, 9.4%, *p*<0.001), antibiotic therapy (n=36, 67.9% vs n=19, 16.2%, *p*<0.001) and antiviral treatments (n=38, 67.9% vs n=2, 1.7%, *p*<0.001) in the COVID-19 group.

Laboratory analysis showed an increased neutrophil and platelet count, reduced lymphocyte count, increased C-reactive protein, erythrocyte sedimentation rate, high-sensitivity troponin T, lactate dehydrogenase, aspartate and alanine aminotransferase, prothrombin time and fibrinogen (all *p*<0.05) in patients with COVID-19 compared to non-COVID-19 patients. No differences were observed for whole white blood cell count, hemoglobin, bilirubin, creatine kinase, creatinine, activated thromboplastin time, d-dimer or serum ferritin (all *p*>0.05) (see **Table 1**).

Patients with COVID-19 had worse functional outcomes as measured by the mRS (5.0, IQR 2.3-6.0 vs 2.0, IQR 1.0-3.0, *p*<0.001), with similar premorbid mRS scores (1.0, IQR 1.0-2.0 vs 1.0, IQR 0.0-2.0, *p*=0.903).

We observed a significant increase in cerebrovascular disease rates in the COVID-19 group (n=43, 76.8% vs n=67, 57.3%, *p*=0.018), with a similar distribution between transient ischemic attack (n=5, 11.6% vs n=8, 11.9%), ischemic stroke (n=35, 81.4% vs n=50, 74.6%) and hemorrhagic stroke (n=3, 7.0% vs n=9, 13.4%) within groups, *p*=0.560 (see **Table 2**).

**Table 2.** Demographic, clinical, laboratory characteristics, treatment and clinical outcomes of patients with cerebrovascular disease.

|  | **Total**<br>**(n=111)** | **non-COVID-19**<br>**(n=68)** | **COVID-19**<br>**(n=43)** | **p-value** |
| --- | --- | --- | --- | --- |
| **Cerebrovascular event** |  |  |  | 0.560 |
| Transient ischemic attack | 13 (11.7%) | 8 (11.8%) | 5 (11.6%) |  |
| Ischemic stroke | 85 (76.6%) | 50 (73.5%) | 35 (81.4%) |  |
| Hemorrhagic stroke | 13 (11.7%) | 10 (14.7%) | 3 (7.0%) |  |
| **Demographic and clinical characteristics** |  |  |  |  |
| Age, years | 76.3 (61.4-82.5) | 74.8 (58.3-80.4) | 76.9 (66.8-85.2) | 0.204 |
| Sex |  |  |  | 0.765 |
| Female | 49 (44.1%) | 28 (41.2%) | 21 (48.8%) |  |
| Male | 62 (55.9%) | 40 (58.8%) | 22 (51.2%) |  |
| Current smoker | 7 (6.3%) | 6 (8.8%) | 1 (2.3%) | 0.526 |
| Comorbidity |  |  |  |  |
| Diabetes | 24 (21.6%) | 14 (20.6%) | 10 (23.3%) | 0.781 |
| Hypercholesterolemia | 33 (29.7%) | 23 (33.8%) | 10 (23.3%) | 0.526 |
| Hypertension | 77 (69.4%) | 51 (75.0%) | 26 (60.5%) | 0.388 |
| Coronary heart disease | 16 (14.4%) | 8 (11.8%) | 8 (18.6%) | 0.643 |
| Malignancy | 24 (21.6%) | 19 (27.9%) | 5 (11.6%) | 0.220 |
| Chronic Kidney Disease | 5 (4.5%) | 3 (4.4%) | 2 (4.7%) | 1.000 |
| Immunodeficiency | 4 (3.6%) | 3 (4.4%) | 1 (2.3%) | 1.000 |
| qSOFA score | 0.7 (0.6-0.8) | 0.5 (0.4-0.7) | 0.9 (0.7-1.1) | 0.077 |
| **Laboratory findings** |  |  |  |  |
| White-cell count (mm<sup>3</sup>) | 8.5 (6.6-10.6) | 8.3 (6.6-10.5) | 8.9 (6.7-11.7) | 0.424 |
| Neutrophil count (mm<sup>3</sup>) | 6.1 (4.5-8.5) | 5.5 (4.1-8.1) | 7.3 (5.0-9.6) | 0.067 |
| Lymphocyte count (mm<sup>3</sup>) | 1.3 (0.9-1.8) | 1.5 (1.2-2.1) | 1.0 (0.8-1.4) | <0.001 |
| Platelet count (mm<sup>3</sup>) | 231.0 (181.0-294.0) | 211.5 (174.5-264.3) | 276.0 (197.0-315.0) | 0.036 |
| Hemoglobin (g/dl) | 13.2 (11.5-14.6) | 12.7 (11.5-14.4) | 13.4 (12.1-14.9) | 0.133 |
| C-reactive protein (mg/L) | 8.1 (2.0-35.7) | 5.1 (1.5-12.9) | 28.0 (4.4-71.1) | <0.001 |
| Erythrocyte sedimentation rate (mm/1h) | 25.0 (14.0-50.0) | 19.5 (12.3-30.8) | 49.0 (29.3-85.0) | <0.001 |
| High-sensitivity troponin T, ng/L | 16.0 (10.0-29.0) | 16.0 (9.3-21.3) | 27.0 (10.0-41.0) | 0.067 |
| Lactate dehydrogenase (U/L) | 220.0 (168.0-277.5) | 177.0 (156.8-221.5) | 277.7 (239.0-365.0) | <0.001 |
| Aspartate aminotransferase (U/L) | 27.7 (21.0-38.0) | 24.0 (21.0-30.5) | 36.0 (28.0-52.0) | <0.001 |
| Alanine aminotransferase (U/L) | 25.0 (16.0-35.0) | 20.5 (15.0-27.8) | 31.0 (20.0-40.7) | <0.001 |
| Total bilirubin (μmol/L) | 0.64 (0.48-0.94) | 0.64 (0.48-0.91) | 0.66 (0.48-1.0) | 0.424 |
| Creatine kinase (U/L) | 109.0 (66.0-241.0) | 104.5 (65.3-221.5) | 114.0 (69.0-337.0) | 0.424 |
| Creatinine (μmol/L) | 0.95 (0.77-1.14) | 0.95 (0.77-1.14) | 0.95 (0.77-1.11) | 0.864 |
| Prothrombin time (sec) | 12.5 (12.0-14.1) | 12.4 (11.8-13.0) | 12.8 (12.2-15.6) | 0.029 |
| Activated thromboplastin time (sec) | 29.0 (26.8-31.0) | 29.1 (26.7-31.4) | 28.6 (27.3-30.7) | 0.770 |
| D-dimer (mg/L) | 693.0 (325.0-2868.0) | 317.0 (197.0-437.0) | 735.5 (364.3-2910.8) | 0.626 |
| Fibrinogen (mg/dL) | 351.0 (291.0-466.0) | 312.0 (261.1-386.4) | 471.0 (368.0-545.0) | <0.001 |
| Serum ferritin (µg/L) | 411.0 (244.0-739.0) | 152.0 (38.0-266.0) | 452.5 (268.0-871.0) | 0.130 |
| **Treatments and exams** |  |  |  |  |
| Antibiotic treatment | 39 (35.1%) | 11 (16.2%) | 28 (65.1%) | <0.001 |
| Antiviral treatment | 27 (24.3%) | 0 (0.0%) | 27 (62.8%) | <0.001 |
| High-flow oxygen therapy | 40 (36.0%) | 7 (10.3%) | 33 (76.7%) | <0.001 |
| Diagnostic tests, n | 6.0 (4.0-8.0) | 6.0 (5.0-8.0) | 6.0 (4.0-8.0) |  |
| Acute phase therapies (ischemic stroke only) | 22 (25.9%) | 13 (26.0%) | 9 (25.7%) | 1.000 |
| Intravenous fibrinolysis | 6 (6.1%) | 2 (3.4%) | 4 (10.0%) | 0.384 |
| Endovascular therapy | 10 (10.2%) | 8 (13.8%) | 2 (5.0%) | 0.785 |
| Bridging therapy | 6 (7.1%) | 3 (6.0%) | 3 (8.6%) | 0.785 |
| **Outcomes** |  |  |  |  |
| In hospital mortality | 19 (17.1%) | 4 (5.9%) | 15 (34.9%) | <0.001 |
| Incident delirium | 13 (11.7%) | 4 (5.9%) | 9 (20.9%) | 0.047 |
| Fever during hospitalization | 29 (26.1%) | 8 (11.8%) | 21 (48.8%) | <0.001 |
| Hospital length of stay, days | 5.0 (4.0-8.0) | 5.0 (4.0-8.0) | 6.0 (4.0-9.0) | 0.425 |
| Modified Rankin Scale pre-morbid | 1.0 (0.0-2.0) | 1.0 (0.0-2.0) | 1.0 (0.0-1.0) | 0.727 |
| Modified Rankin Scale discharge | 2.0 (1.0-5.0) | 2.0 (1.0-3.0) | 5.0 (2.0-6.0) | <0.001 |
| Good outcome | 59 (53.2%) | 48 (70.6%) | 11 (25.6%) | <0.001 |
| NIH Stroke Scale admission | 5.0 (2.0-15.0) | 4.0 (2.0-14.8) | 10.0 (3.0-15.0) | 0.147 |
| NIH Stroke Scale discharge | 3.0 (0.0-10.0) | 2.0 (0.0-6.8) | 9.0 (1.0-19.0) | 0.005 |

Data are median (IQR), or n (%). *p*-values were calculated by Mann-Whitney U test, $\chi^2$ test or Fisher's exact test, as appropriate, corrected for multiple comparisons using the Benjamini-Hochberg false discovery rate (FDR).
qSOFA=quick Sequential Organ Failure Assessment; combined therapy=intravenous fibrinolysis followed by endovascular therapy

Patients admitted for ischemic stroke had similar baseline characteristics, including sex, comorbidities, premorbid mRS and NIHSS on admission. There were no differences in access to acute phase therapies as endovascular treatment (n=2, 5.0% vs n=8, 13.8%, *p*=0.785), intravenous fibrinolysis (n=4, 10.0% vs n=2, 3.4%, *p*=0.384) or bridging therapy (n=3, 8.6% vs n=3, 6.0%, *p*=0.785). Patients with COVID-19 had higher mRS scores at discharge (5.0, IQR 2.0-6.0 vs 2.0, IQR 1.0-3.0, *p*<0.001), with a significantly lower number of patients with a good outcome (n=11, 25.6% vs n=48, 70.6%, *p*<0.001). This difference in outcomes was also confirmed considering only patients who underwent acute phase therapies (mRS score at discharge *p*=0.009; good outcome *p*=0.047).

Moreover, patients with cerebrovascular disease (including transient ischemic attack, ischemic and hemorrhagic stroke) and COVID-19 had an increased platelet count, reduced lymphocyte count, higher C-reactive protein, erythrocyte sedimentation rate, lactate dehydrogenase, aspartate and alanine aminotransferase, prothrombin time and fibrinogen levels compared to patients with cerebrovascular disease but without COVID-19 (see **Table 2**).

In univariable analysis, increased age, higher qSOFA scores, thrombocytopenia, elevated C-reactive protein and lactate dehydrogenase were associated with in-hospital death in the COVID-19 group (**Table 3**). In the multivariable logistic regression model, we found that higher qSOFA scores (odds ratio 4.47, 95% CI 1.21-16.50; *p*=0.025), lower platelets (0.98, 0.97-0.99; *p*=0.005) and higher lactate dehydrogenase (1.01, 1.00-1.03; *p*=0.009) on admission were all associated with increased odds of death in COVID-19 patients (**Table 3**).

**Table 3.**
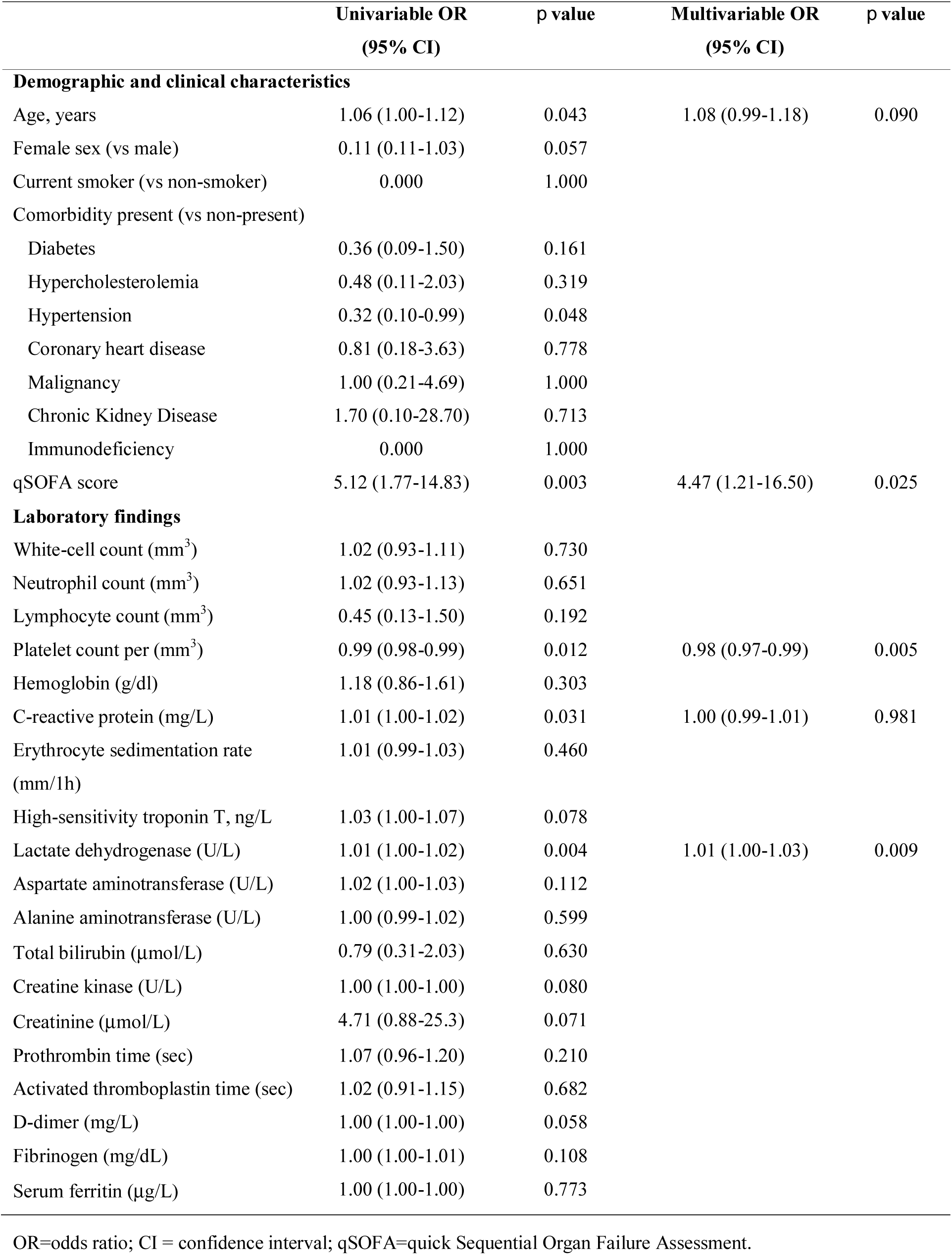
Risk factors on admission associated with in-hospital mortality in COVID-19 patients.

|  | Univariable OR<br>(95% CI) | *p* value | Multivariable OR<br>(95% CI) | *p* value |
| --- | --- | --- | --- | --- |
| **Demographic and clinical characteristics** |  |  |  |  |
| Age, years | 1.06 (1.00-1.12) | 0.043 | 1.08 (0.99-1.18) | 0.090 |
| Female sex (vs male) | 0.11 (0.11-1.03) | 0.057 |  |  |
| Current smoker (vs non-smoker) | 0.000 | 1.000 |  |  |
| Comorbidity present (vs non-present) |  |  |  |  |
| Diabetes | 0.36 (0.09-1.50) | 0.161 |  |  |
| Hypercholesterolemia | 0.48 (0.11-2.03) | 0.319 |  |  |
| Hypertension | 0.32 (0.10-0.99) | 0.048 |  |  |
| Coronary heart disease | 0.81 (0.18-3.63) | 0.778 |  |  |
| Malignancy | 1.00 (0.21-4.69) | 1.000 |  |  |
| Chronic Kidney Disease | 1.70 (0.10-28.70) | 0.713 |  |  |
| Immunodeficiency | 0.000 | 1.000 |  |  |
| qSOFA score | 5.12 (1.77-14.83) | 0.003 | 4.47 (1.21-16.50) | 0.025 |
| **Laboratory findings** |  |  |  |  |
| White-cell count (mm<sup>3</sup>) | 1.02 (0.93-1.11) | 0.730 |  |  |
| Neutrophil count (mm<sup>3</sup>) | 1.02 (0.93-1.13) | 0.651 |  |  |
| Lymphocyte count (mm<sup>3</sup>) | 0.45 (0.13-1.50) | 0.192 |  |  |
| Platelet count per (mm<sup>3</sup>) | 0.99 (0.98-0.99) | 0.012 | 0.98 (0.97-0.99) | 0.005 |
| Hemoglobin (g/dl) | 1.18 (0.86-1.61) | 0.303 |  |  |
| C-reactive protein (mg/L) | 1.01 (1.00-1.02) | 0.031 | 1.00 (0.99-1.01) | 0.981 |
| Erythrocyte sedimentation rate<br>(mm/1h) | 1.01 (0.99-1.03) | 0.460 |  |  |
| High-sensitivity troponin T, ng/L | 1.03 (1.00-1.07) | 0.078 |  |  |
| Lactate dehydrogenase (U/L) | 1.01 (1.00-1.02) | 0.004 | 1.01 (1.00-1.03) | 0.009 |
| Aspartate aminotransferase (U/L) | 1.02 (1.00-1.03) | 0.112 |  |  |
| Alanine aminotransferase (U/L) | 1.00 (0.99-1.02) | 0.599 |  |  |
| Total bilirubin (μmol/L) | 0.79 (0.31-2.03) | 0.630 |  |  |
| Creatine kinase (U/L) | 1.00 (1.00-1.00) | 0.080 |  |  |
| Creatinine (μmol/L) | 4.71 (0.88-25.3) | 0.071 |  |  |
| Prothrombin time (sec) | 1.07 (0.96-1.20) | 0.210 |  |  |
| Activated thromboplastin time (sec) | 1.02 (0.91-1.15) | 0.682 |  |  |
| D-dimer (mg/L) | 1.00 (1.00-1.00) | 0.058 |  |  |
| Fibrinogen (mg/dL) | 1.00 (1.00-1.01) | 0.108 |  |  |
| Serum ferritin (μg/L) | 1.00 (1.00-1.00) | 0.773 |  |  |

OR=odds ratio; CI = confidence interval; qSOFA=quick Sequential Organ Failure Assessment.

## Discussion

This retrospective cohort study identified several differences between patients with and without COVID-19 who were hospitalized for a neurological disease. In particular, patients with COVID-19 were older, had higher qSOFA scores on admission, and had a higher rate of cerebrovascular disorders compared to patients without COVID-19. During hospitalization, COVID-19 patients had a higher incidence of delirium and fever, with prolonged hospital length of stay and increased in-hospital mortality rates. Additionally, patients with COVID-19 had significant differences in laboratory values on admission, including blood count analysis, acute phase proteins and coagulation profiles.

We identified potential risk factors for a poor prognosis at an early stage, as high qSOFA score, thrombocytopenia and increased lactate dehydrogenase levels. Previous reports have also found that SOFA and qSOFA scores were associated with in-hospital mortality, as well as d-dimer and older age, in adult inpatients with COVID-19.^3^ The qSOFA is a bedside prompt that may identify patients with suspected infection who are at greater risk for a poor outcome outside the intensive care unit, and may be rapidly performed by the clinician without the need of laboratory analysis.^12^

Interestingly, we observed a significant increase of stroke rates in patients with COVID-19, with worse outcomes compared to the non-COVID-19 group, including higher mRS scores at discharge and a significantly lower number of patients with a good outcome, at par with access to acute phase therapies. As recently suggested in a statement by the American Heart Association and by the American Stroke Association (AHA/ASA) Stroke Council Leadership, stroke mechanisms in COVID-19 could include different processes, including the release of pro-inflammatory cytokines with a direct effect on plaque rupture through local inflammation and activation of coagulation factors, or cardioembolism from virus-related cardiac injury.^13–17^

Moreover, a direct effect of the virus on endothelial cells or on heart tissue could be hypothesized, considering that the receptor for SARS-CoV-2, the angiotensin converting enzyme 2,^18–20^ is expressed on vascular endothelial cells and myocytes.^21–23^

In our cohort we observed several indexes of an altered coagulability in patients with COVID-19 compared to patients without COVID-19. Prothrombin time and d-dimer were increased in the former group, including inflammatory indexes as C-reactive protein and erythrocyte sedimentation rate. This marker profile is consistent with what has been observed in disseminated intravascular coagulation and may play an important role in stroke incidence and severity in COVID-19 patients.^24^ Abnormal coagulation parameters have been also shown to be associated with poor prognosis in patients with COVID-19 associated pneumonia.^25^

From the present analysis, patients who underwent endovascular therapy or intravenous fibrinolysis in the COVID-19 group had more severe outcomes, including increase in NIHSS and mRS scores at discharge. This raises a critical issue if patients with COVID-19 should be equally treated with acute phase therapies; further studies on broader populations should try to shed further light on this matter.

What also emerges from this study is that differences in laboratory and most clinical features between neurology admissions of patients with and without COVID-19 mostly reflect the features of COVID-19 infection, including fever, thrombocytopenia, elevated lactate dehydrogenase, and high morbidity and mortality.^3,9,26,27^ The concomitant neurological comorbidity, as stroke or other major neurological disorder, could possibly increase even further the high mortality observed in this patients group. Moreover, it is still not clear if COVID-19 may increase the incidence of stroke^28^ and other neurological diseases, as encephalitis^29^ or immune-mediated neurological disorders.^30–32^ With the surge of intensive care unit admissions during the pandemic,^2^ and as underlined by the AHA/ASA Stroke Council Leadership, it has become necessary to prioritize intensive care unit resources, at the possible expenses of stroke patients,^15,33^ withholding ventilation when necessary and raising issues on potential legal liabilities.^34^ These aspects should all be addressed in neurology units that deal with acute phase diseases and COVID-19 patients.

We acknowledge that this study entails some limitations. First, due to the retrospective study design, not all laboratory tests were performed in all patients, including high sensitivity troponin T, ferritin and d-dimer, therefore their role could have not been thoroughly assessed in this study. Second, results on stroke do not take into account several factors due to the retrospective design, including stroke subtypes, infarct volume and recanalization rates. Third, interpretation of our findings could be limited by sample size and by the single-center design. Fourth, there could be a selection bias due to the unwillingness of patients with COVID-related symptoms or infection to come for hospital neurological evaluation unless extremely necessary, as with stroke, epilepsy or other major neurological disorders. Further larger, multi-center, prospective studies should be performed to confirm these findings.

To the best of our knowledge, this is the largest retrospective cohort study among patients with neurological disorders and COVID-19, with a definite outcome. Patients with COVID-19 and neurological disease have worse clinical and neurological outcomes, with higher in-hospital mortality rates compared to patients without COVID-19.

## Data Availability

All study data, including raw and analysed data, and materials will be available from the corresponding author, A.P., upon reasonable request.

## Acknowledgments

We would like to express our gratitude to the nurses, auxiliary staff, technicians and all the colleagues who collaborated for the management of patients during the COVID-19 outbreak. Moreover, we would like to thank Dr. Marco Trivelli, Dr. Camilo Rossi and Dr. Loretta Jacquot for their support in setting up a Neuro-COVID Unit.

## Declaration of interests

The authors declare no competing interests.

## Notes

### Competing Interest Statement

A. Benussi, A. Pilotto, E. Premi, I. Libri, M. Giunta, C. Agosti, A. Alberici, E. Baldelli, M. Benini, S. Bonacina, L. Brambilla, S. Caratozzolo, M. Cortinovis, A. Costa, S. Cotti Piccinelli, E. Cottini, V. Cristillo, I. Delrio, M. Gamba, S. Gazzina, N. Gilberti, S. Gipponi, A. Imarisio, P. Invernizzi, U. Leggio, M. Leonardi, P. Liberini, M. Locatelli, S. Masciocchi, L. Poli, R. Rao, B. Risi, L. Rozzini, A. Scalvini, F. Schiano di Cola, R. Spezi, V. Vergani, I. Volonghi, N. Zoppi, B. Borroni, M. Magoni, A. Pezzini report no disclosures relevant to the manuscript. A. Padovani is consultant and served on the scientific advisory board of GE Healthcare, Eli-Lilly and Actelion Ltd Pharmaceuticals, received speaker honoraria from Nutricia, PIAM, Lansgstone Technology, GE Healthcare, Lilly, UCB Pharma and Chiesi Pharmaceuticals.

### Funding Statement

No external funding was received.

